# Novel Representations of Vaccine Protection Against Progression to Severe Disease Over Time

**DOI:** 10.64898/2026.02.12.26346197

**Authors:** Natalie E Dean, Veronika I. Zarnitsyna

**Affiliations:** Department of Biostatistics & Bioinformatics, Emory Rollins School of Public Health; Department of Microbiology & Immunology, Emory School of Medicine

## Abstract

**Background:** Vaccines can prevent severe disease by preventing infection or by reducing progression among those who become infected. Vaccine effectiveness against progression given infection is often used to quantify this second mechanism, but it conditions on infection, which is itself affected by vaccination. As a result, this estimand lacks a clear causal interpretation and may behave non-intuitively over time.

**Methods:** We introduce a conceptual framework that models protection against infection and protection against progression as separate components that wane over time. Protection is represented using individual-level threshold-crossing times that depend on covariates and define a time-varying population susceptible to infection. Within this framework, we derive standard vaccine effectiveness estimands and propose two alternative decompositions of protection against severe disease: a progression-risk–weighted multiplicative decomposition and an additive decomposition based on absolute risk reduction. We illustrate their behavior using simulated examples.

**Results:** The weighted multiplicative decomposition restores a causal interpretation for progression protection within the doomed principal stratum and avoids negative estimates. The additive decomposition provides a clear representation of the pathways over time.

**Conclusions:** Explicitly modeling the infection-to-severe-disease pathway improves interpretation of vaccine effectiveness under waning immunity.

## 1. Introduction

Vaccines can prevent severe disease outcomes through at least two mechanisms: by preventing infection entirely, or by reducing the risk of progression to severe disease following infection. The latter mechanism is often more durable, reflecting immune responses that act later in the disease course and wane more slowly than those that block infection (1-3). As a result, many vaccines retain substantial protection against severe disease even after protection against infection has declined (3-8).

The fact that vaccines can protect against severe disease even when breakthrough infection occurs has posed a communication challenge, including for COVID-19 vaccines. The CDC’s messaging campaign that “a flu vaccine can take flu from wild to mild” communicates this well (9). While estimates of vaccine effectiveness against severe disease support this message, it is noteworthy that we lack well-defined and interpretable effectiveness measures that specifically target protection against progression.

Vaccine effectiveness against progression to disease given infection (*VE*_*P*_) is the primary estimand available (10), but it has limitations. This estimand compares outcomes among infected vaccinated and infected unvaccinated individuals, but vaccination itself affects who becomes infected. As a result, the infected vaccinated group is typically enriched for individuals with weaker immune protection or greater underlying vulnerability, as evidenced by their breakthrough status. Consequently, *VE*_*P*_ lacks a causal interpretation. Some causal inference approaches, such as principal stratification methods developed for post-infection outcomes in vaccine trials, address some aspects of this problem by targeting latent subgroups (11, 12). However, these approaches typically do not incorporate time-varying loss of protection, which is central to understanding vaccine performance in real-world settings.

The conditional estimand *VE*_*P*_ is special in that it serves as a bridge between the unconditional estimands of vaccine effectiveness against infection (*VE*_*S*_) and vaccine effectiveness against severe disease (*VE*_*SP*_) (10). Prior work showed that *VE*_*P*_ can exhibit strange behavior, including increasing over time since vaccination, that is unlikely to reflect real strengthening of second-line immune protection (13). Instead, the increase most likely reflects changes in the composition of the infected population over time, further reducing the measure’s real-world interpretability.

In this paper, we introduce a new approach for characterizing the progression benefit of a vaccine. We define a novel conceptual framework that explicitly models protection against infection and protection against progression as functions of time since vaccination. This framework naturally defines a population susceptible to infection that changes over time. Building on this structure, we show how commonly reported VE estimands relate to one another over time. In response, we propose a novel multiplicative decomposition and a novel additive decomposition of protection against severe disease, each yielding a more interpretable summary of vaccine effects under waning immunity. We demonstrate the added interpretability with a series of simulated examples.

## 2. Conceptual Framework: A Generalization of the All-or-Nothing Model

We conceptualize vaccine protection as operating at two distinct points along the disease pathway: (1) susceptibility to infection, and (2) progression to severe disease following infection. We model the immune protection that underlies each step as separate components, which we refer to as “first-line” and “second-line” protection, respectively. In this framework, there are two ways to prevent severe disease: by preventing infection or progression (see **Figure 1**).

**Figure 1.**
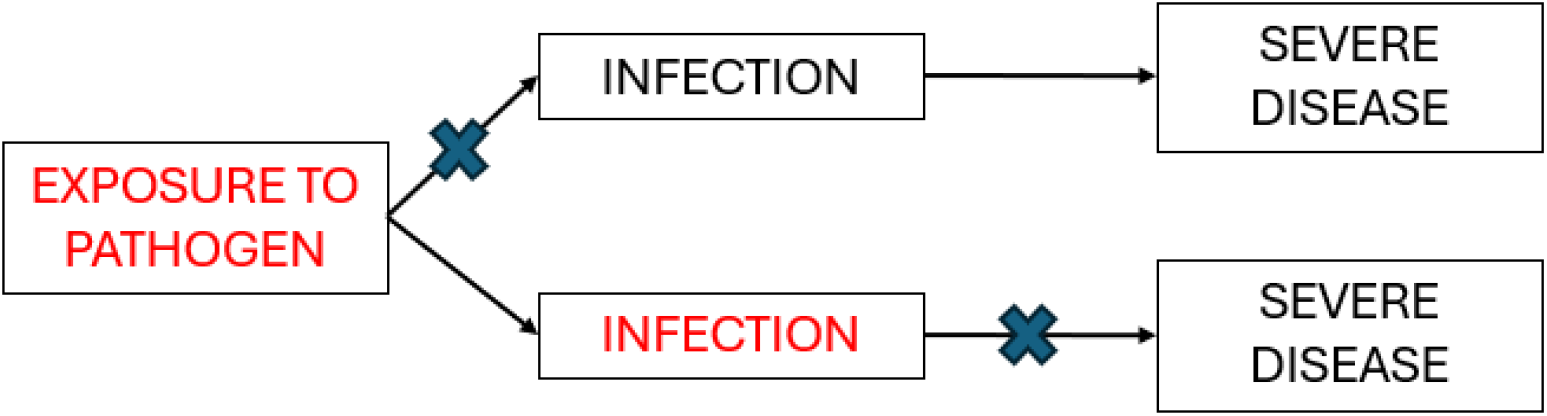
Conceptual framework for protection against infection and progression to severe disease. Vaccination provides protection at two points along the disease pathway: first-line protection against infection and second-line protection against progression to severe disease following infection. Severe disease can be prevented either by averting infection or, among infected individuals, by reducing the risk of progression.

We assume that vaccine-conferred first- and second-line protection reach an initial peak after vaccination that subsequently wanes over time. While we do not specify the exact form of this trajectory, we assume that there exists a time at which each protection threshold is crossed. We also allow that, for some individuals, protection never exceeds the threshold, meaning they are never vaccine-protected.

Let denote the time since vaccination at which first-line protection drops below a threshold (i.e., protection against infection is lost). Here, represents individual-level covariates that influence the durability of an individual’s vaccine protection, such as age or presence of comorbidities. Before time *T*_l_(*X*), an individual is not susceptible to infection (full protection) because their protection is above the threshold. After *T*_l_(*X*), the individual loses all protection against infection and is considered “infectable” if exposed. Thus, for first-line protection, we draw inspiration from the classic all-or-nothing vaccine model (14). Importantly, though, we generalize this model to allow protection to *wane over time* rather than being a fixed characteristic, and we allow for population heterogeneity through *X*.

To model progression, we start by defining a baseline (i.e., unvaccinated) risk of progression to severe disease given infection, *π* (*X*). This risk depends on covariates and allows for heterogeneity in underlying vulnerability, such as elevated risk among infants, older adults, or immunocompromised individuals. Second-line vaccine protection then operates to reduce this probability by a multiplicative factor *g*(·). Like with first-line protection, we assume each vaccinated person has a threshold time *T*_2_(*X*), before which they are fully protected against progression to severe disease, i.e., the multiplicative factor is 0 for *t* ≤ *T*_2_(*X*). For times *t* > *T*_2_(*X*), protection begins to wane according to the multiplier *g*(*t* − *T*_2_,*X*), expressed as a function of time since crossing the threshold (*t* − *T*_2_(*X*)) and covariates *X*. For example, this could wane linearly with some slope that depends upon *X*. We leave the form general, although we specify a special case of all-or-nothing protection for progression, i.e., multiplier *g*(*t* − *T*_2_(*X*),*X*) for all *t* > *T*_2_(*X*), meaning no remaining protection.

## 3. Standard VE Estimands Over Time Under the Proposed Framework

Under the framework described in **Section 2**, we derive three time-varying population-level protection functions corresponding to commonly reported vaccine effectiveness estimands (10).

First, we define vaccine effectiveness against susceptibility to infection, *VE*_*S*_ (*t*). This is one minus the relative risk of infection if everyone vaccinated time units ago is exposed versus if everyone unvaccinated is exposed, assuming no indirect transmission effects. We assume no prior protection, so all unvaccinated are susceptible to infection. For simplicity, we assume the same exposure and infection risk across all levels of *X*, but we describe how to relax this assumption in the **Supplement**. Under the all-or-nothing representation for infection susceptibility, we show in the **Supplement** that *VE*_*S*_ (*t*) is given by:

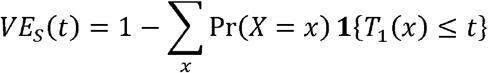

*VE*_*S*_ (*t*) is thus the fraction of vaccinated individuals who have not yet crossed the threshold *T*_l_(*X*) for loss of protection against infection by time *t* since vaccination. Thus, *T*_l_(*X*) naturally defines a time-varying set of individuals susceptible to infection.

Second, we define vaccine effectiveness against severe disease, *VE*_*SP*_ (*t*). This summarizes protection against experiencing severe disease by time *t*, and we show in the Supplement that it can be written as:

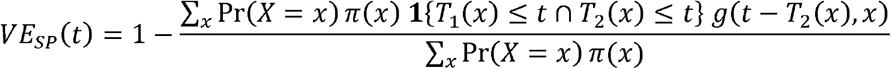

The numerator is the expected proportion with severe disease if all vaccinated susceptible are infected at time *t* after vaccination. The denominator is the expected proportion with severe disease if all unvaccinated are infected, i.e., *E*_*x*_ [*π* (*X*)]. Unlike *VE*_*S*_ (*t*), *VE*_*SP*_ (*t*) is weighted by baseline progression risk *π* (*X*) (see **Supplement**).

Finally, vaccine effectiveness against progression to severe disease given infection, *VE*_*P*_(*t*), is the connective tissue between *VE*_*S*_ (*t*) and *VE*_*SP*_ (*t*), as seen by their relationship:

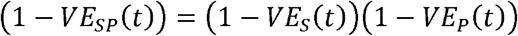

In our framework, *VE*_*P*_(*t*) can be written as follows (see **Supplement**):

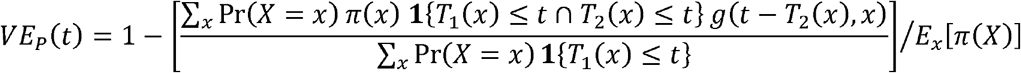

Within the brackets is the average progression risk of the *susceptible* vaccinated at time *t*. This is compared to the average progression risk if all are unvaccinated and infected.

Unlike the unconditional measures *VE*_*S*_ (*t*) and *VE*_*SP*_ (*t*), vaccine effectiveness against progression, *VE*_*P*_(*t*), conditions on infection, or here susceptibility to infection defined by having *T*_l_(*X*) ≤ *t* or being unvaccinated. As long as *VE*_*S*_ (*t*) > 0, the susceptible vaccinated population at time *t* will be a subset of the overall vaccinated population, and often this subset has a covariate profile *X* corresponding to weaker second-line protection and/or higher baseline progression risk. As observed previously, *VE*_*P*_(*t*) can exhibit strange behavior, such as being negative or increasing over time, making it challenging to interpret (13).

## 4. An Alternative Multiplicative Decomposition of Protection Against Severe Disease

We propose a novel decomposition of *VE*_*SP*_ (*t*) that instead uses versions of the above estimates weighted by baseline severe disease risk:

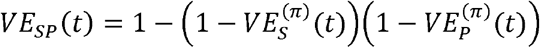

As shown in the **Supplement**, this decomposition is built upon two new estimands:

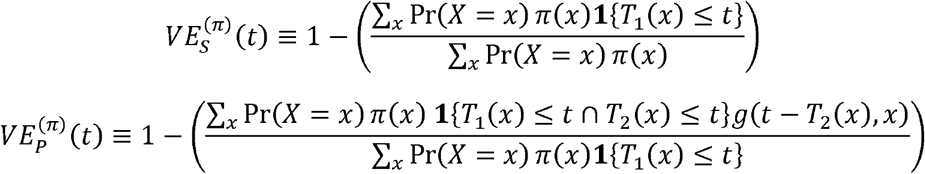

The term 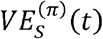 is a weighted version of *VE*_*S*_ (*t*), with weights according to the baseline risk of severe disease *π* (*X*). This captures the relative reduction in severe disease attributable to <u>infection prevention</u>. This is not “infections prevented,” but rather “severe cases prevented by averting infections,” which in turn depends on who is at risk risk of severe disease, i.e., *π* (*X*). The term 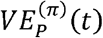 captures the reduction in progression among those who are susceptible to infection at time *t* regardless of vaccination status, defined by *T*_l_(*x*) ≤ *t*. Another name for this subgroup is the <u>“doomed” principal stratum</u> at time *t* (see **Supplement**).

Using the “doomed” principal stratum is appealing as the progression term now has a causal interpretation. Yet its interpretation is not straightforward when we study changes in 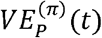 over time. Notably, membership in the “doomed” principal stratum is growing over time as more people lose infection protection.

This is best illustrated by decomposing the denominator of 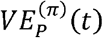 into two terms: (1) those who retain full protection against severe disease, and (2) those who do not:

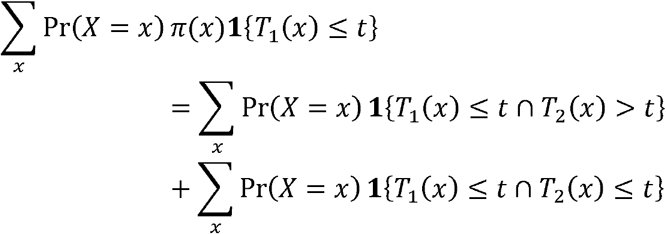

As shown in **Figure 2**, while we may be primarily interested in how individuals within the doomed principal stratum lose protection (i.e., move from the middle group to the right), there is a competing force as more individuals enter the doomed principal stratum at each time step (i.e., move from the left group to the middle).

**Figure 2.**
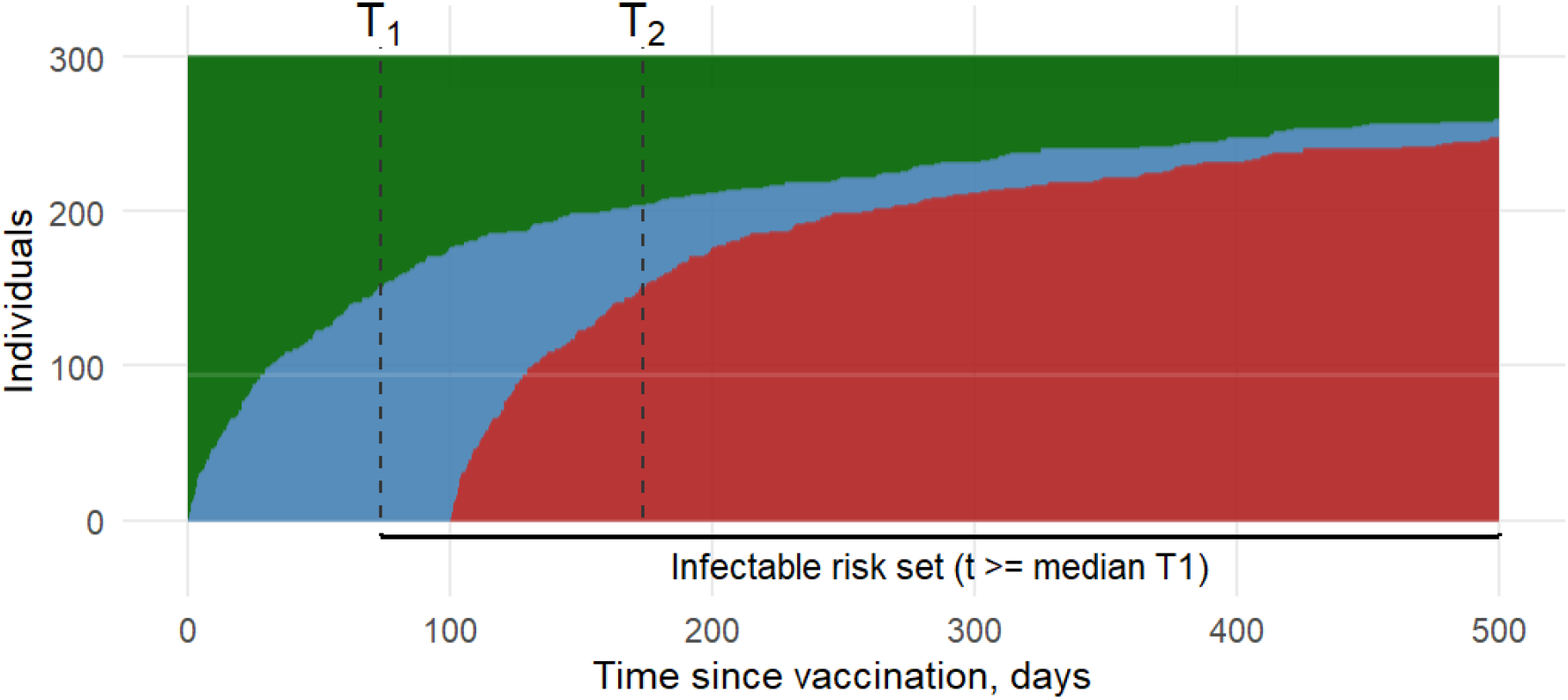
Threshold-crossing representation of waning first- and second-line protection. This plot shows the individual trajectories of 300 hypothetical individuals, stacked horizontally and sorted by their crossing time *T*_l_. In this example, we set *T*_2_ = *T*_l_ + 100. The green compartment is the time when vaccinated individuals are protected against infection (*t* ≤ *T*_l_(*X*)). The blue compartment is the time when vaccinated individuals are susceptible to infection but protected against severe disease (*T*_l_(*X*) < *t* ≤ *T*_2_(*X*)). The red compartment is the time when vaccinated individuals are susceptible and at risk of severe disease (*t* > *T*_2_(*X*)). The doomed population at time *t* is the sum of the blue and red compartments. The inflow into the doomed population drives non-intuitive behavior of progression-based VE measures.

Thus, we note a close relationship between 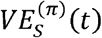 and 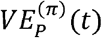, with individuals only contributing to the progression measure once they have lost their initial protection.

## 5. An Additive Decomposition of Protection Against Severe Disease

To offer greater interpretability, we further propose an additive decomposition. Under our framework, we show in the **Supplement** that *VE*_*SP*_ (*t*) can be decomposed by adding two prevention pathways:

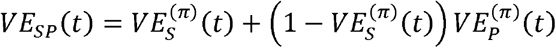

As before, 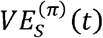 can be interpreted as the reduction in severe disease risk attributable to infection prevention. The second term captures the reduction in severe disease risk attributable to preventing progression, driven by our new term 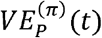 but *scaled* by 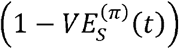. We can see how the second term quantifying prevention of progression can only grow after protection against infection wanes.

## 6. Illustrative Examples

We explore the behavior of the existing decomposition and the proposed decompositions through a series of illustrative examples. Full details are provided in the **Supplement**. Briefly, we model a hypothetical population with five subgroups denoted by the covariate *X* ∈ −2, −1,0,1,2. Subgroups with higher values of *X* have poorer health, denoted by higher baseline progression risk *π* (*X*), e.g., *π* (*X*) = 0.01,0.02,0.05,0.15,0.30 with population mean 0.10. We draw *T*_l_(*X*) from an exponential proportional hazards model with a mean *µ*_l_ (e.g., 100 days) and positive slope *β*_l_ for the effect of *X*. This induces highest hazard and shortest *T*_l_ for highest values of *X*. We use the same procedure to draw *T*_2_(*X*), with mean *µ*_2_ (e.g., 300 days) and positive slope *β*_2_. *T*_l_(*X*) and *T*_2_cx) are then marginally correlated when *β*_l_ and *β*_2_ are non-zero. Given simulated *T*_l_ and *T*_2_ for 200,000 individuals, we can calculate and plot each of the decompositions.

In the first example, we consider the case where *T*_l_(*x*) = *T*_2_(*X*) under all-or-nothing second-line protection. We generate this case by setting *T*_2_ to equal *T*_l_ for all individuals. In this setting, vaccine <u>only</u> prevents infection, and once infected, the risk of severe disease returns to the baseline value *π* (*X*). Intuitively, one might expect that *VE*_*S*_ (*t*) = *VE*_*SP*_ (*t*) in this case, implying *VE*_*P*_(*t*). However, in most realistic settings, individuals who lose protection first will also have higher baseline risk *π* (*X*) than the general population. The mean baseline risk among the doomed population changes as a function of time *t*, starting above the population average of 10% (see **Supplement**).

Over time, it approaches the population average as eventually all vaccinated enter the doomed population.

As shown in **Figure 3**, the poor infection prevention in high risk individuals yields 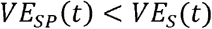. For 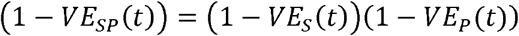 to hold, *VE*_*P*_(*t*) < 0. This example reveals that one important role of *VE*_*P*_(*t*) as “connective tissue” can be to <u>discount</u> a vaccine with poor performance in those most vulnerable to severe disease. In contrast, in our proposed multiplicative decomposition, 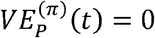 and there is no progression component in the additive decomposition.

**Figure 3.**
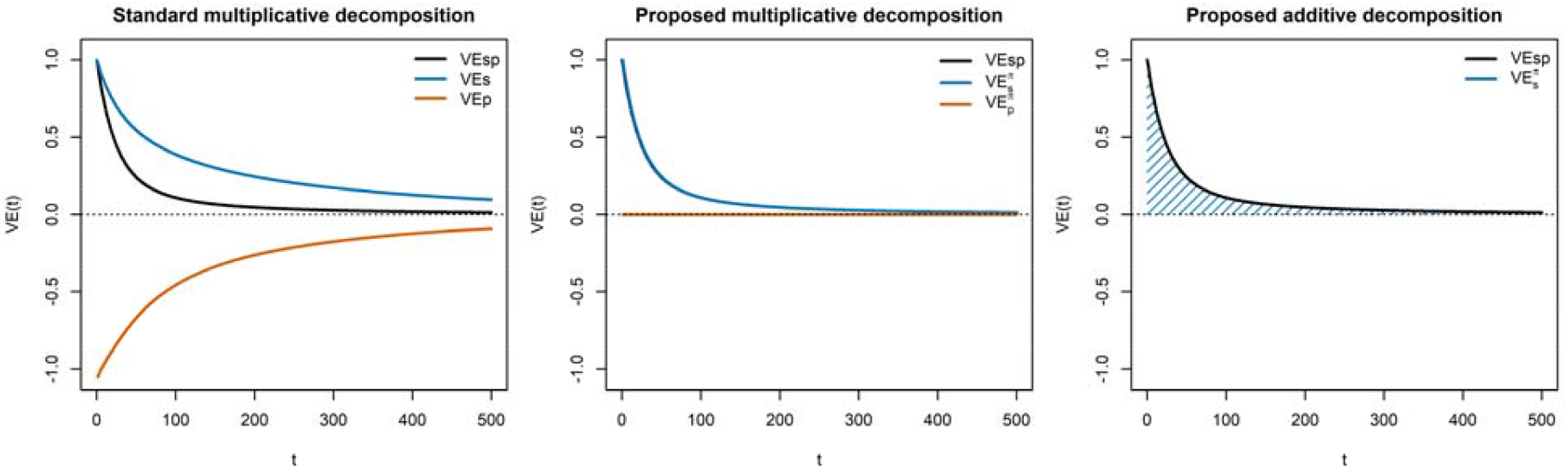
Standard and proposed decompositions of protection against severe disease when loss of infection and progression protection coincide. *Illustrative example in which the time to loss of protection against infection and progression are identical (T*_l_ *= T*_2_*), so the vaccine confers no additional protection against progression beyond prevention of infection. Left panel: Standard multiplicative decomposition of vaccine effectiveness against severe disease (VE*_*SP*_*) into effectiveness against infection (VE*_*S*_*) and effectiveness against progression given infection (VE*_*P*_*), which yields VE*_*S*_ *> VE*_*SP*_ *and non-intuitive negative VE*_P_. *Middle panel: Proposed progression-risk–weighted multiplicative decomposition, in which VE*_*SP*_ *factors into a weighted infection component* 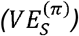 *and a progression component defined within the doomed principal stratum* 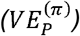; *here* 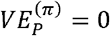 *as expected. Right: Proposed additive decomposition of the absolute reduction in severe disease risk, showing that all protection is attributed to prevention of infection when T*_l_ *= T*_2_.

In **Figure 4**, we provide a second example where *T*_l_(*X*) ≠ *T*_l_(*X*) but where *T*_l_(*X*), *T*_2_(*X*), and *π* (*X*) are marginally correlated via the shared covariate *X*. Like in the prior example, *VE*_*SP*_ (*t*) < *VE*_*S*_ (*t*)for some because the vaccine does relatively worse at preventing infection in those at highest risk of severe disease. When considering those who are susceptible to infection, the susceptible vaccinated have <u>higher baseline risk</u> *π* (*X*) than the population average *E*_*X*_[*π* (*X*)], leading to negative *VE*_*P*_(*t*). This is due to selection induced by heterogeneity in *T*_l_(*X*). In contrast, in the proposed multiplicative decomposition, 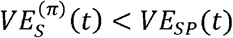. Furthermore, the additive decomposition offers a visual representation of how these mechanisms split over time.

**Figure 4.**
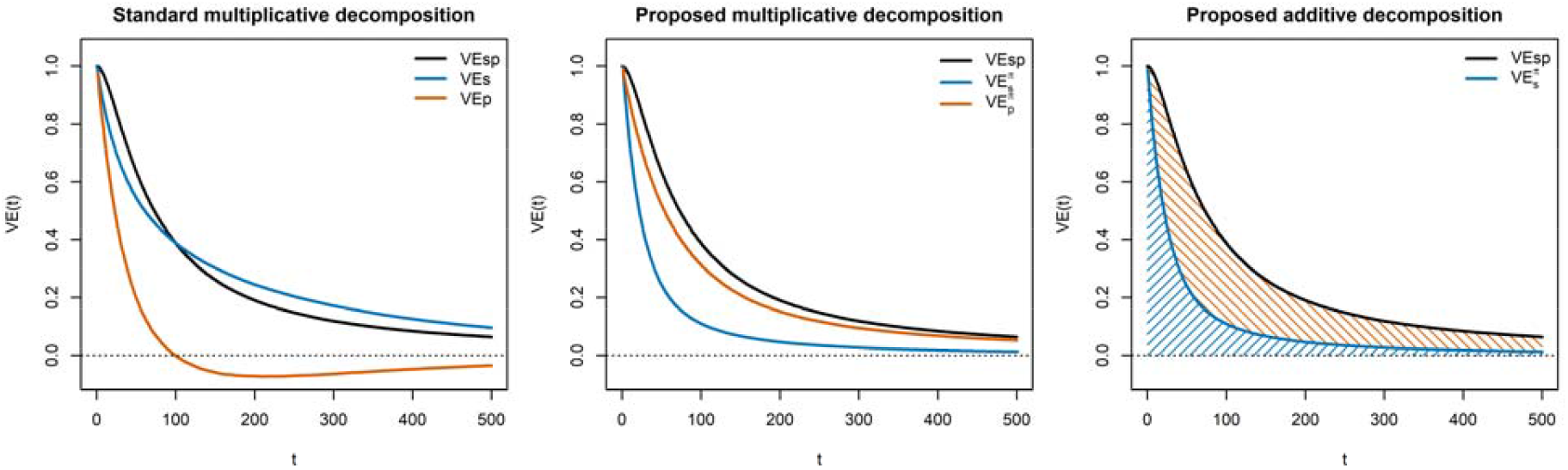
Standard and proposed decompositions of protection against severe disease with more durable progression protection. Illustrative example in which protection against progression to severe disease wanes more slowly than protection against infection (*µ*_l_ *=* 100, *µ*_2_ = 300). *Left panel:* Standard multiplicative decomposition of vaccine effectiveness against severe disease (*VE*_*SP*_) into effectiveness against infection (*VE*_*S*_) and effectiveness against progression given infection (*VE*_*P*_). As protection against infection wanes, *VE*_P_ exhibits non-intuitive behavior driven by changes in the composition of the susceptible vaccinated. *Middle panel:* Proposed progression-risk–weighted multiplicative decomposition. This decomposition yields components with clearer interpretation over time. *Right panel:* Proposed additive decomposition of the absolute reduction in severe disease risk, showing separate contributions from prevention of infection and prevention of progression among infected individuals. As protection against infection wanes, the contribution of progression protection naturally grows, making the source of protection against severe disease easy to see.

To better understand the system, we identify conditions when 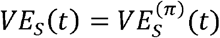. When these are equal, 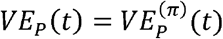 since each pair must multiply to *VE*_*SP*_ (*t*). This occurs when either of two conditions is met: (1) there is no effect modification in infection protection, i.e., *T*_l_(*X*) does not depend upon *X*, and/or (2) there is no heterogeneity in baseline progression risk, i.e., *π* (*X*) ≡ *π* across individuals. When either of these is true, then there is no mechanism selecting vulnerable individuals into the doomed group early on.

Next, we consider fixing 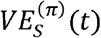 and explore major drivers of 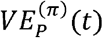 and, in turn, *VE*_*SP*_ (*t*). In our base scenario where *T*_l_, *T*_2_, and *π* depend upon *X* and so are correlated, *VE*_*SP*_ (*t*) can be very low. This occurs because the vulnerable individuals are also those with the shortest protection against severe disease. The performance of the vaccine is very sensitive to the durability of protection in the most vulnerable individuals, as these individuals contribute to the majority of severe disease cases. In **Figure 5**, we observe that 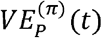 and *VE*_*SP*_ (*t*) can increase substantially by reducing the association between *T*_2_ and *X*, even keeping the marginal means the same. This is achieved by setting the slope *β*_2_ to a smaller value or even 0.

**Figure 5.**
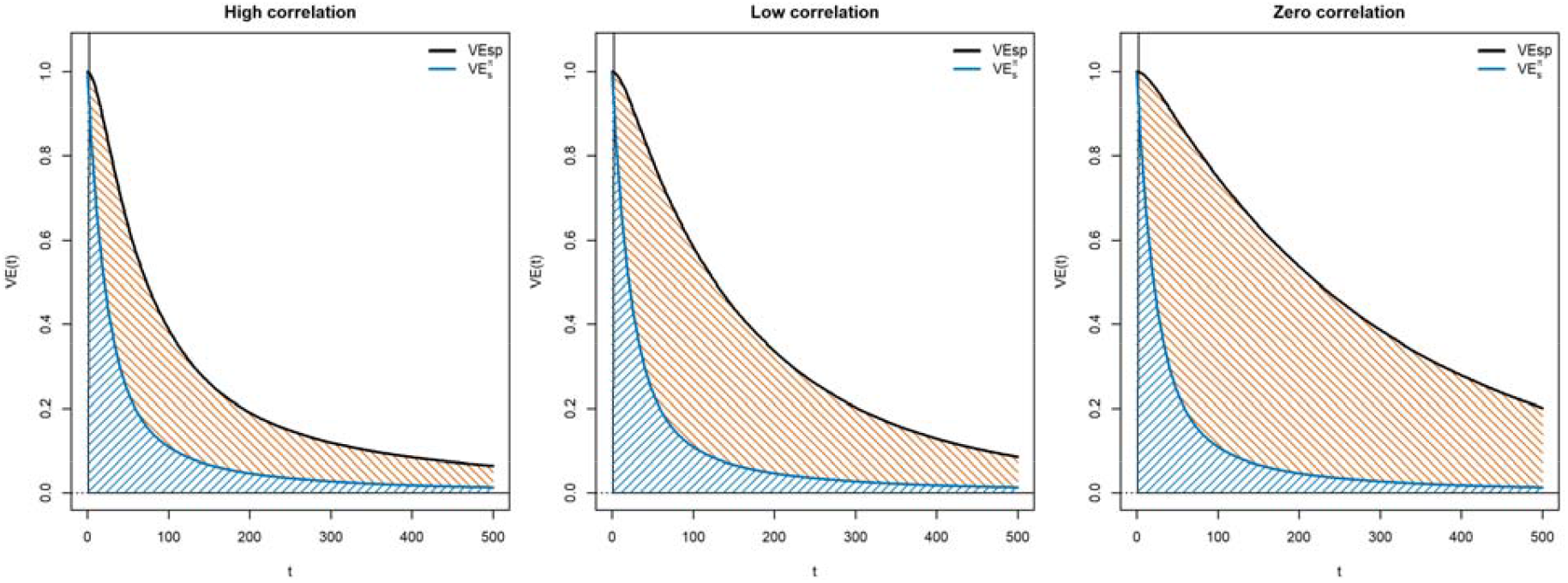
Additive decomposition under varying correlation between first- and second-line protection. Illustrative examples showing the additive decomposition of protection against severe disease under high, low, and zero correlation between loss of infection protection and loss of progression protection. The high-correlation setting (left) is the base scenario with *β*_2_ = 0.8. The low-correlation setting (middle) reduces this parameter to *β*_2_ = 0.4, and the zero-correlation setting (right) sets *β*_2_ = 0. All settings have the same median *T*_2_ value. Shaded regions represent the contributions of infection prevention (blue) and progression prevention among infected individuals (orange) to the total effectiveness against severe disease (*VE*_SP_, black). As correlation decreases, a larger share of protection against severe disease is attributed to progression prevention.

## 7. Extension to Leaky Protection

In our primary framework, we represent vaccine-induced protection using threshold-crossing times, which make the evolution of the susceptible population explicit. However, the phenomena we describe are not artifacts of all-or-nothing protection. The same selection mechanisms arise under fully leaky protection models in which vaccination continuously reduces, but does not eliminate, susceptibility to infection and risk of progression following infection. In a leaky formulation, vaccinated individuals retain a positive probability of infection and disease at all times, with these risks varying across individuals and waning over time.

In this setting, conditioning on infection reweights the vaccinated population toward individuals with higher residual susceptibility or weaker immune responses. When heterogeneity in infection protection is correlated with baseline progression risk or with the durability of second-line protection, vaccine effectiveness against progression given infection can exhibit time-varying, non-monotonic, or even negative behavior driven by changes in risk-set composition rather than changes in progression biology. Thus, these interpretability challenges arise generally from conditioning on post-vaccination infection under heterogeneous waning, not from the threshold formulation itself.

Importantly, the additive decomposition of protection against severe disease extends naturally to leaky protection models and does not rely on threshold-based definitions of infectability. When severe disease risk factorizes multiplicatively into infection and progression components, the absolute reduction in severe disease can be decomposed into severe cases averted through infection prevention and severe cases averted through progression prevention among infections that still occur. This remains true no matter how vaccine protection is modeled: whether it completely blocks risk or only partially reduces it. We describe this in greater detail in the **Supplement**.

## 8. Discussion

Vaccines can prevent severe disease by preventing infection and by reducing progression to severe illness among those who become infected. In this paper, we developed a framework that makes this structure explicit over time. By representing protection as threshold-crossing times *T*_l_(*X*) and *T*_2_(*X*), we created a time-varying subset susceptible to infection. Within this framework, we derived both multiplicative and additive decompositions of protection against severe disease that improve interpretability relative to standard measures.

Our results help explain why vaccine effectiveness against progression, *VE*_*P*_(*t*), can behave in non-intuitive ways. First, *VE*_*S*_ (*t*) and *VE*_*SP*_ (*t*) are defined over different weightings of the population. Protection against severe disease is inherently weighted by baseline progression risk, whereas protection against infection is not. As a result, *VE*_*S*_ (*t*) can theoretically exceed *VE*_*SP*_ (*t*) when infection prevention is concentrated among lower-risk individuals, yielding a negative *VE*_*P*_(*t*) . Second, *VE*_*P*_(*t*) is defined within a time-varying group whose composition changes as protection against infection wanes. When loss of infection protection varies across covariate strata, conditioning on infection induces selection. These features disappear under restrictive assumptions, such as constant baseline progression risk or no effect modification in *T*_l_(*X*), but such conditions are unlikely to hold in practice.

We proposed a novel progression-risk–weighted multiplicative decomposition of *VE*_*SP*_ (*t*) that restores a clearer causal interpretation to the progression term. In this representation, protection against infection is weighted by baseline severe disease risk, and the progression component is the relative risk within the doomed principal stratum. This eliminates pathological behavior, such as a negative progression measure. However, even in this formulation, the progression term is difficult to interpret over time as the doomed stratum expands. Increases in *VE*_*P*_(*t*) can reflect changes in population composition rather than strengthening biological protection.

For this reason, we also proposed an additive decomposition of *VE*_*SP*_ (*t*), derived from the absolute risk reduction. This representation separates the contribution of infection prevention from progression prevention in terms of the number of severe cases averted. As protection against infection wanes, the relative contribution of progression prevention naturally increases, even if second-line protection remains stable. The additive framework makes this shift transparent and avoids conflating changes in population composition with changes in biological efficacy. We believe that this representation may be especially useful for communication and surveillance reporting.

Our findings connect to broader epidemiologic phenomena such as depletion of susceptibles and frailty selection. The non-intuitive behavior of progression-based estimands arises from conditioning on a subset whose risk profile changes over time. What may appear as strengthening or weakening of protection can instead reflect evolving composition of the infected group. Making the structure explicit helps distinguish biological mechanisms from selection effects.

This work has several limitations. We assumed homogeneous exposure and baseline infection risk across individuals and did not allow exposure to vary over time since vaccination. In practice, exposure may differ across covariate strata and may change with behavior or risk compensation (15), which would introduce additional weighting and could further shape the relative contributions of infection and progression prevention. We also did not explicitly model prior infection in vaccinated or unvaccinated individuals. Depletion of susceptibles over time has an important influence on what is estimated in vaccine studies (16). We believe that representing protection as threshold-crossing times provides a natural path for incorporating infection history, and this is an area for future work. We note that we observed a wider dynamic range in *VE*_*P*_(*t*) from prior modeling (13), which we attribute to depletion of susceptibles that does not occur in this framework. Finally, this paper focused on clarifying estimands and interpretation rather than fully addressing identification or estimation. We can turn to recent developments in causal inference tools for vaccine studies (17). Intuitively, the key additional ingredient for estimation is baseline severe disease risk, which is often observable in surveillance data, but formal estimation strategies remain an important area for future work.

Vaccines protect against severe disease through multiple mechanisms, and those mechanisms wane over time and vary across subpopulations. Standard progression-based measures can have non-intuitive behavior when protection wanes heterogeneously. By decomposing protection using progression-based-weights, we design more interpretable estimands for evaluating vaccine performance against severe outcomes.

## Data Availability

Code available upon request.

