## Supplement for "Novel Representations of Vaccine Protection Against Progression to Severe Disease Over Time"

***SUPPLEMENTARY APPENDIX***

Natalie E Dean^1^, Veronika Zarnitsyna^2^

^1^Department of Biostatistics & Bioinformatics, Emory Rollins School of Public Health

^2^Department of Immunology and Microbiology, Emory School of Medicine

### S1. Notation and Setup

- Covariate $X$, e.g., age groupings
- $T_{1}(X)$, time since vaccination at which first-line protection is lost, assuming all-or-nothing protection against infection. A random variable that depends upon $X$.
- $\pi(X)$, baseline (unvaccinated) risk of progression to severe disease given infection for covariate level $X$.
- $T_{2}(X)$, time since vaccination at which full second-line protection is lost. A random variable that depends upon $X$.
- $g(t-T_{2},X)$, function describing remaining protection against progression to severe disease after $T_{2}(X)$ as a multiplicative factor of $\pi(X)$

Under this model, time is divided into up to three periods. In the first period, there is no risk of infection. In the second period, there is no risk of progression. In the third period, risk of progression is reduced by the multiplier function. In general, $T_{1}\left( x \right)<T_{2}(x)$, but our framework is general. If $T_{2}\left( x \right)<T_{1}(x)$, time is only in two periods. **Figure S1** shows these settings.

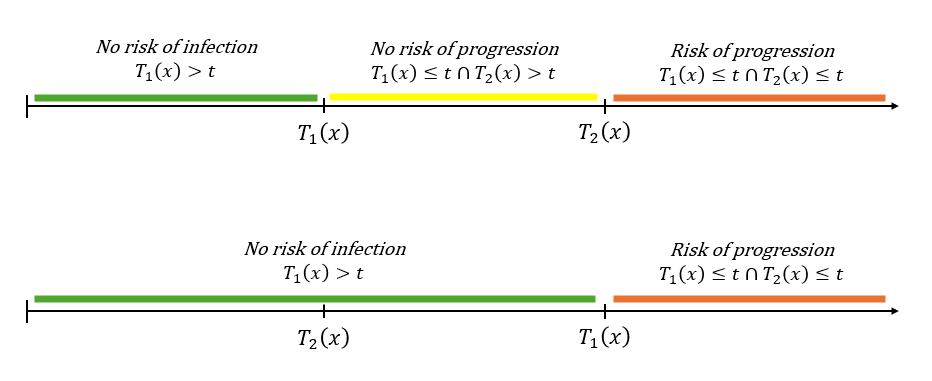

Figure S1. Ordering of loss of infection and progression protection.
Illustration of two possible orderings of the individual-specific threshold crossing times $T_{1}(x)$ and $T_{2}(x)$. In the common setting (top), protection against infection is lost before protection against progression ($T_{1}(x)<T_{2}(x)$), creating an intermediate period in which individuals are infectable but remain protected against severe disease. The lower panel shows the reverse ordering ($T_{1}(x)>T_{2}(x)$), in which progression protection is lost first. Although this latter case is expected to be rare in practice, our framework and derivations allow for either ordering.

### S2. Derivation of Standard VE Relationships

Protection against susceptibility to infection:

The denominator assumes all unvaccinated are susceptible to infection.

$$VE_{S}\left( t \right)=1-\frac{\sum_{x} \Pr\left( X=x \right)\boldsymbol{1}\left\{ T_{1}\left( x \right)\leq t \right\}}{\sum_{x} \Pr\left( X=x \right)}$$

$$=\sum_{x} \Pr\left( X=x \right)\boldsymbol{1}\left\{ T_{1}\left( x \right)>t \right\}$$

Protection against severe disease:

The denominator assumes that all unvaccinated are susceptible to infection.

$$VE_{SP}\left( t \right)=1-\frac{\sum_{x} \Pr\left( X=x \right)\pi\left( x \right)\boldsymbol{1}\left\{ T_{1}\left( x \right)\leq t\cap T_{2}\left( x \right)\leq t \right\}g(t-T_{2},X)}{\sum_{x} \Pr\left( X=x \right)\pi(x)}$$

Note that $\boldsymbol{1}\left\{ T_{1}\left( x \right)\leq t\cap T_{2}\left( x \right)\leq t \right\}$ captures the population that has lost both first- and second-line protection. It gives more weight to the groups that have higher $\pi(x)$. The weights here are the *fraction of severe cases that would occur in this group if no one in the population was vaccinated*. In the extreme, a group with zero (or very low) $\pi(x)$ will have essentially no impact on the waning of $VE_{SP}(t)$.

Protection against progression:

$$1-VE_{P}\left( t \right)=\left[ \frac{\sum_{x} \Pr\left( X=x \right)\pi\left( x \right)\boldsymbol{1}\left\{ T_{1}\left( x \right)\leq t\cap T_{2}\left( x \right)\leq t \right\}g(t-T_{2},X)}{\sum_{x} \Pr\left( X=x \right)\boldsymbol{1}\left\{ T_{1}\left( x \right)\leq t \right\}} \right]/{\sum_{x} \Pr\left( X=x \right)\pi(x)}$$

$$=\left[ \frac{\sum_{x} \Pr\left( X=x \right)\pi\left( x \right)\boldsymbol{1}\left\{ T_{1}\left( x \right)\leq t\cap T_{2}\left( x \right)\leq t \right\}g(t-T_{2},X)}{\left( 1-VE_{S}\left( t \right) \right)} \right]/{E_{x}\left[ \pi\left( X \right) \right]}$$

### S3. Multiplicative Decomposition of $\boldsymbol{V}\boldsymbol{E}_{\boldsymbol{SP}}\boldsymbol{(t)}$

We derive an alternative multiplicative decomposition of $VE_{SP}(t)$, which we derive here:

$$1-VE_{SP}\left( t \right)=\frac{\sum_{x} \Pr\left( X=x \right)\pi\left( x \right)\boldsymbol{1}\left\{ T_{1}\left( x \right)\leq t\cap T_{2}\left( x \right)\leq t \right\}g\left( t-T_{2}\left( x \right),x \right)}{\sum_{x} \Pr\left( X=x \right)\pi(x)}$$

$$=\frac{\sum_{x} \Pr\left( X=x \right)\pi\left( x \right)\boldsymbol{1}\left\{ T_{1}\left( x \right)\leq t\cap T_{2}\left( x \right)\leq t \right\}g\left( t-T_{2}\left( x \right),x \right)}{\sum_{x} \Pr\left( X=x \right)\pi(x)}\left( \frac{\sum_{x} \Pr\left( X=x \right)\pi\left( x \right)\boldsymbol{1}\left\{ T_{1}\left( x \right)\leq t \right\}}{\sum_{x} \Pr\left( X=x \right)\pi\left( x \right)\boldsymbol{1}\left\{ T_{1}\left( x \right)\leq t \right\}} \right)$$

Multiply and divide by the same term

$$=\left( \frac{\sum_{x} \Pr\left( X=x \right)\pi\left( x \right)\boldsymbol{1}\left\{ T_{1}\left( x \right)\leq t \right\}}{\sum_{x} \Pr\left( X=x \right)\pi\left( x \right)} \right)\left( \frac{\sum_{x} \Pr\left( X=x \right)\pi\left( x \right)\boldsymbol{1}\left\{ T_{1}\left( x \right)\leq t\cap T_{2}\left( x \right)\leq t \right\}g\left( t-T_{2}\left( x \right),x \right)}{\sum_{x} \Pr\left( X=x \right)\pi\left( x \right)\boldsymbol{1}\left\{ T_{1}\left( x \right)\leq t \right\}} \right)$$

Regroup

$$\equiv\left( 1-VE_{S}^{\left( \pi\right)}\left( t \right) \right)\left( 1-VE_{P}^{\left( \pi\right)}\left( t \right) \right)$$

We note the alignment between $VE_{P}^{\left( \pi\right)}\left( t \right)$ and the “doomed” vaccine susceptible stratum at time $t$. These are individuals who would be susceptible by time $t$, regardless of vaccination, defined by $T_{1}\left( x \right)\leq t$.

| **Principal stratum** | **Outcome if exposed at** $\boldsymbol{t}$**, assuming vaccinated at 0** | **Outcome if exposed at** $\boldsymbol{t}$**, assuming never vaccinated** |
| --- | --- | --- |
| Always infected (Doomed) | Infected | Infected |
| Vaccine protected | Prevented | Infected |
| Never infected | Prevented | Prevented |
| Vaccine harmed (assume empty) | Infected | Prevented |

We can show that the relative risk of progression in the “doomed” stratum at time $t$ is equal to $1-VE_{P}^{\left( \pi\right)}\left( t \right)$:

$$\frac{\left[ \frac{\sum_{x} \Pr\left( X=x \right)\pi\left( x \right)\boldsymbol{1}\left\{ T_{1}\left( x \right)\leq t\cap T_{2}\left( x \right)\leq t \right\} g\left( t-T_{2}\left( x \right),x \right)}{\sum_{x} \Pr\left( X=x \right)\boldsymbol{1}\left\{ T_{1}\left( x \right)\leq t \right\}} \right]}{\left[ \frac{\sum_{x} \Pr\left( X=x \right)\pi\left( x \right)\boldsymbol{1}\left\{ T_{1}\left( x \right)\leq t\cap T_{2}\left( x \right)\leq t \right\}}{\sum_{x} \Pr\left( X=x \right)\boldsymbol{1}\left\{ T_{1}\left( x \right)\leq t \right\}} \right]}$$

$$=\left[ \frac{\sum_{x} \Pr\left( X=x \right)\pi\left( x \right)\boldsymbol{1}\left\{ T_{1}\left( x \right)\leq t\cap T_{2}\left( x \right)\leq t \right\} g\left( t-T_{2}\left( x \right),x \right)}{\sum_{x} \Pr\left( X=x \right) \pi(x)\boldsymbol{1}\left\{ T_{1}\left( x \right)\leq t \right\}} \right]$$

$$=1-VE_{P}^{\left( \pi\right)}\left( t \right)$$

### S4. Additive Decomposition of $\boldsymbol{V}\boldsymbol{E}_{\boldsymbol{SP}}\boldsymbol{(t)}$

We start by defining absolute risk reduction of severe disease $ARR_{SP}(t)$. This is the absolute reduction in severe disease cases if everyone is unvaccinated and exposed to infection, minus if everyone is vaccinated and exposed to infection, assuming no indirect effects.

$$ARR_{SP}\left( t \right)=\sum_{x} \Pr\left( X=x \right)\pi\left( x \right)-\sum_{x} \Pr\left( X=x \right)\pi\left( x \right)\boldsymbol{1}\left\{ T_{1}\left( x \right)\leq t\cap T_{2}\left( x \right)\leq t \right\}g\left( t-T_{2}\left( x \right),x \right)$$

$$=\sum_{x} \Pr\left( X=x \right)\pi\left( x \right)\left[ 1-\frac{\sum_{x} \Pr\left( X=x \right)\pi\left( x \right)\boldsymbol{1}\left\{ T_{1}\left( x \right)\leq t\cap T_{2}\left( x \right)\leq t \right\}g\left( t-T_{2}\left( x \right),x \right)}{\sum_{x} \Pr\left( X=x \right)\pi\left( x \right)} \right]$$

$$=E_{x}\left[ \pi\left( X \right) \right] VE_{SP}\left( t \right)$$

We note that $ARR_{SP}(t)$ can be written as the product of mean baseline risk $E_{x}[\pi\left( X \right)]$ and vaccine protection against severe disease $VE_{SP}(t)$.

To derive a new decomposition, we can return to the first line and add/subtract an intermediate term:

$$ARR_{SP}\left( t \right)=\sum_{x} \Pr\left( X=x \right)\pi\left( x \right)-\sum_{x} \Pr\left( X=x \right)\pi\left( x \right)\boldsymbol{1}\left\{ T_{1}\left( x \right)\leq t \right\}+\sum_{x} \Pr\left( X=x \right)\pi\left( x \right)\boldsymbol{1}\left\{ T_{1}\left( x \right)\leq t \right\}-\sum_{x} \Pr\left( X=x \right)\pi\left( x \right)\boldsymbol{1}\left\{ T_{1}\left( x \right)\leq t\cap T_{2}\left( x \right)\leq t \right\}g\left( t-T_{2}\left( x \right),x \right)$$

We group the first two terms and simplify:

$$\left( first two terms \right)=\sum_{x} \Pr\left( X=x \right)\pi\left( x \right)-\sum_{x} \Pr\left( X=x \right)\pi\left( x \right)\boldsymbol{1}\left\{ T_{1}\left( x \right)\leq t \right\}$$

$$=\sum_{x} \Pr\left( X=x \right)\pi\left( x \right)\left( 1-\frac{\sum_{x} \Pr\left( X=x \right)\pi\left( x \right)\boldsymbol{1}\left\{ T_{1}\left( x \right)\leq t \right\}}{\sum_{x} \Pr\left( X=x \right)\pi\left( x \right)} \right)$$

$$=E_{x}\left[ \pi\left( X \right) \right] VE_{S}^{\left( \pi\right)}(t)$$

We group the last two terms and simplify:

$$\left( last two terms \right)=\sum_{x} \Pr\left( X=x \right)\pi\left( x \right)\boldsymbol{1}\left\{ T_{1}\left( x \right)\leq t \right\}-\sum_{x} \Pr\left( X=x \right)\pi\left( x \right)\boldsymbol{1}\left\{ T_{1}\left( x \right)\leq t\cap T_{2}\left( x \right)\leq t \right\}g\left( t-T_{2}\left( x \right),x \right)$$

Divide both terms by $\sum_{x} \Pr\left( X=x \right)\pi\left( x \right)\boldsymbol{1}\left\{ T_{1}\left( x \right)\leq t \right\}$ and factor this out.

$$=\left( \sum_{x} \Pr\left( X=x \right)\pi\left( x \right)\boldsymbol{1}\left\{ T_{1}\left( x \right)\leq t \right\} \right)\left( 1-\frac{\sum_{x} \Pr\left( X=x \right)\pi\left( x \right)\boldsymbol{1}\left\{ T_{1}\left( x \right)\leq t\cap T_{2}\left( x \right)\leq t \right\}g\left( t-T_{2}\left( x \right),x \right)}{\sum_{x} \Pr\left( X=x \right)\pi\left( x \right)\boldsymbol{1}\left\{ T_{1}\left( x \right)\leq t \right\}} \right)$$

Multiply and divide by $\sum_{x} \Pr\left( X=x \right)\pi\left( x \right)$. Replace last term with $VE_{P}^{\left( \pi\right)}(t)$:

$$=\left( \sum_{x} \Pr\left( X=x \right)\pi\left( x \right) \right)\left( \frac{\sum_{x} \Pr\left( X=x \right)\pi\left( x \right)\boldsymbol{1}\left\{ T_{1}\left( x \right)\leq t \right\}}{\sum_{x} \Pr\left( X=x \right)\pi\left( x \right)} \right) VE_{P}^{\left( \pi\right)}\left( t \right)$$

$$=E_{x}\left[ \pi\left( X \right) \right]\left( 1-VE_{S}^{\left( \pi\right)}\left( t \right) \right)VE_{P}^{\left( \pi\right)}\left( t \right)$$

Thus, we have shown that:

$$E_{x}\left[ \pi\left( X \right) \right] VE_{SP}\left( t \right)=E_{x}\left[ \pi\left( X \right) \right] VE_{S}^{\left( \pi\right)}\left( t \right)+E_{x}\left[ \pi\left( X \right) \right]\left( 1-VE_{S}^{\left( \pi\right)}\left( t \right) \right)VE_{P}^{\left( \pi\right)}\left( t \right)$$

We divide everything by $E_{x}\left[ \pi\left( X \right) \right]$ to return our decomposition:

$$VE_{SP}\left( t \right)= VE_{S}^{\left( \pi\right)}\left( t \right)+\left( 1-VE_{S}^{\left( \pi\right)}\left( t \right) \right)VE_{P}^{\left( \pi\right)}\left( t \right)$$

### S5. Simulation Setup

We simulated vaccinated populations of size $n$with heterogeneity in baseline risk of severe disease indexed by a discrete covariate $X\in\{-2,-1,0,1,2\}$, equally distributed across individuals. Baseline risk of progression given infection, $\pi(X)$, was specified either as constant (10% for all groups) or varying across $X$ with fixed mean 10% (values 0.01, 0.02, 0.05, 0.12, 0.30). First-line protection (protection against infection) and second-line protection (protection against progression) were represented by individual-specific threshold times $T_{1}$and $T_{2}$, respectively. These were generated independently from exponential distributions with proportional hazards structure:

$$\lambda_{1}(X)=\lambda_{10}\exp(\beta_{1}X),\lambda_{2}(X)=\lambda_{20}\exp(\beta_{2}X),$$

where $\lambda_{10}=1/\text{mean}T1_{X=0}$ and $\lambda_{20}=1/\text{mean}T2_{X=0}$. Varying $\beta_{1}$and $\beta_{2}$ controlled the degree of alignment (correlation) between waning of infection and progression protection. Additional scenarios imposed deterministic relationships (e.g., $T_{2}=T_{1}$ or $T_{2}=T_{1}+100$) to represent perfect or near-perfect overlap. We assumed all-or-nothing protection at first- and second-stage.

### S6. Illustrative Example with equal $\boldsymbol{T}_{\boldsymbol{1}}$, $\boldsymbol{T}_{\boldsymbol{2}}$

If $T_{1}\left( x \right)=T_{2}(x)$ with all-or-nothing protection against severe disease, we can simplify $VE_{SP}(t)$ to:

$$1-VE_{SP}\left( t \right)=\frac{\sum_{x} \Pr\left( X=x \right)\pi\left( x \right)\boldsymbol{1}\left\{ T_{1}\left( x \right)\leq t \right\}}{\sum_{x} \Pr\left( X=x \right)\pi(x)}$$

We note that this is not equal to $VE_{S}(t)$, below, because $VE_{S}(t)$ lacks weighting by $\pi\left( X \right)$:

$$1-VE_{S}\left( t \right)=\sum_{x} \Pr\left( X=x \right)\boldsymbol{1}\left\{ T_{1}\left( x \right)\leq t \right\}$$

We write out $VE_{P}(t)$ in this example:

$$1-VE_{P}\left( t \right)=\left[ \frac{\sum_{x} \Pr\left( X=x \right)\pi\left( x \right)\boldsymbol{1}\left\{ T_{1}\left( x \right)\leq t \right\}}{\sum_{x} \Pr\left( X=x \right)\boldsymbol{1}\left\{ T_{1}\left( x \right)\leq t \right\}} \right]/{\sum_{x} \Pr\left( X=x \right)\pi(x)}$$

This compares the average risk of severe disease progression among the susceptible vaccinated (those who have lost protection by time $t$) with the average risk of severe disease progression in the entire population. Where those who have lost protection by time $t$ have higher risk $\pi(X)$, $VE_{P}(t)$ will be negative.

We can reconsider the $T_{1}\left( X \right)=T_{2}(X)$ example using our proposed multiplicative decomposition. First, we note that $VE_{SP}\left( t \right)=VE_{S}^{\left( \pi\right)}(t)$. Second, we can see that $VE_{P}^{\left( \pi\right)}\left( t \right)=0$:

$$1-VE_{P}^{\left( \pi\right)}\left( t \right)=1-\left( \frac{\sum_{x} \Pr\left( X=x \right)\pi\left( x \right)\boldsymbol{1}\left\{ T_{1}\left( x \right)\leq t \right\}}{\sum_{x} \Pr\left( X=x \right)\pi\left( x \right)\boldsymbol{1}\left\{ T_{1}\left( x \right)\leq t \right\}} \right)$$

### S7. Additional Simulation Results

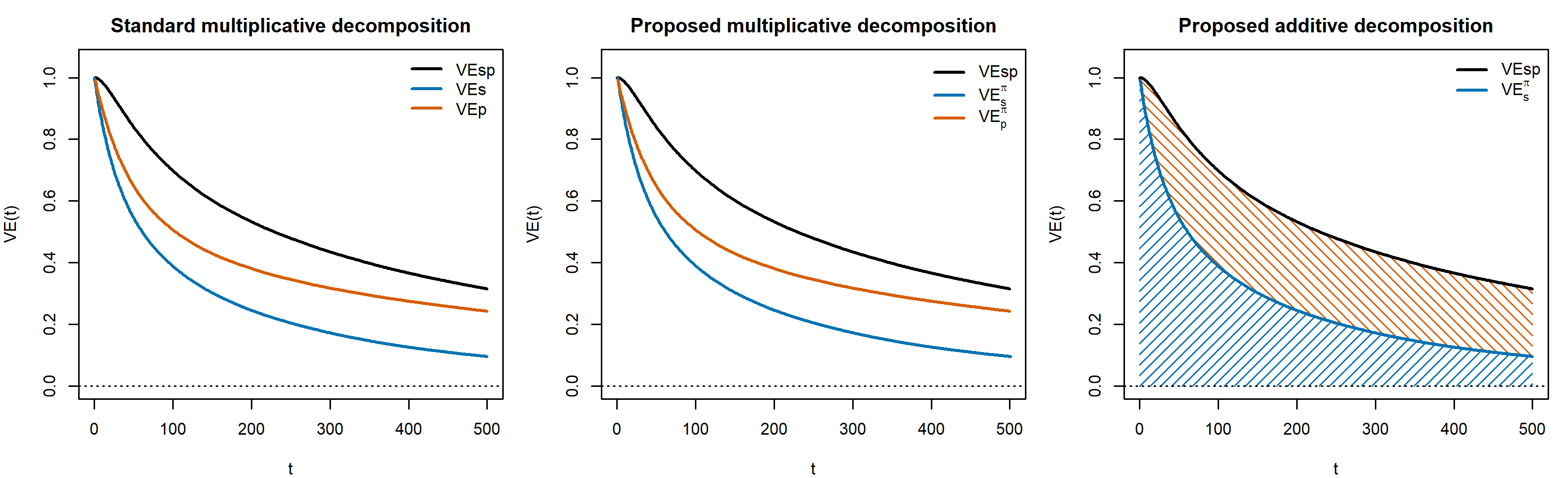

Figure S2. Decompositions when baseline progression risk is constant. Illustrative example in which baseline progression risk $\pi(x)$ is constant across individuals. In this setting, the weighted and unweighted multiplicative decompositions coincide, and the proposed and standard versions yield identical results.

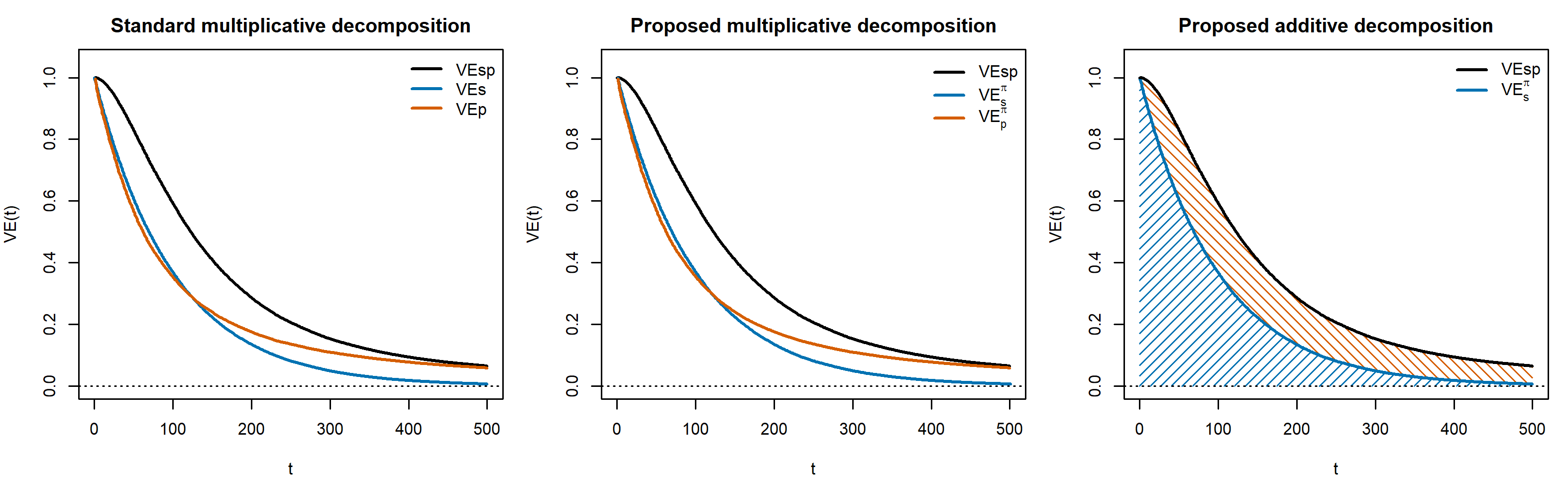

Figure S3. Decompositions when $T_{1}$ does not depend on $X$. Illustrative example in which $T_{1}$ does not depend on $X$, achieved by setting $\beta_{1}=0$. In this setting, the weighted and unweighted multiplicative decompositions coincide, and the proposed and standard versions yield identical results.

### S8. Extension for Variable Exposure and Baseline Infection Risk

We describe how the model extends when exposure and baseline infection risk is variable across the population. We assume that each group has $r(x)$ which is a relative exposure and baseline infection risk multiplier. This captures the fact that some groups have increased exposure, e.g., due to behavioral factors, but also may have different baseline infection risk. We do not attempt to distinguish these but rather summarize them in a single term. Furthermore, we leave extending $r(x)$ to be a function of time since vaccination $t$ for future work.

$$VE_{S}\left( t \right)=1-\frac{\sum_{x} \Pr\left( X=x \right)r(x)\boldsymbol{1}\left\{ T_{1}\left( x \right)\leq t \right\}}{\sum_{x} \Pr\left( X=x \right)r(x)}$$

$$VE_{SP}\left( t \right)=1-\frac{\sum_{x} \Pr\left( X=x \right)r\left( x \right) \pi\left( x \right)\boldsymbol{1}\left\{ T_{1}\left( x \right)\leq t\cap T_{2}\left( x \right)\leq t \right\}g(t-T_{2},X)}{\sum_{x} \Pr\left( X=x \right) r(x)\pi(x)}$$

We note that the groups are now weighted by the combined effect of their exposure/baseline infection risk $r\left( x \right)$ and their baseline progression risk $\pi(x)$.

$$VE_{P}\left( t \right)=1-\left[ \frac{\sum_{x} \Pr\left( X=x \right)r\left( x \right) \pi\left( x \right)\boldsymbol{1}\left\{ T_{1}\left( x \right)\leq t\cap T_{2}\left( x \right)\leq t \right\}g(t-T_{2},X)}{\sum_{x} \Pr\left( X=x \right)r\left( x \right)\boldsymbol{1}\left\{ T_{1}\left( x \right)\leq t \right\}} \right]/{\sum_{x} \Pr\left( X=x \right)r\left( x \right) \pi(x)}$$

We can decompose $VE_{SP}(t)$ in the same way, by multiplying and dividing by $\sum_{x} \Pr\left( X=x \right)\pi\left( x \right) r(x)\boldsymbol{1}\left\{ T_{1}\left( x \right)\leq t \right\}.$

S8. Extension for Leaky Protection

While the main framework summarizes vaccine-induced protection using threshold-crossing times, the same selection mechanisms arise under leaky protection models in which vaccination continuously reduces, but does not eliminate, susceptibility to infection and risk of disease following infection. In a leaky formulation, vaccinated individuals retain a positive probability of infection and disease at all times, with these risks varying across individuals and waning over time.

Let $\lambda_{0}(X)$ denote baseline infection risk under a common exposure regime, and let $S(t\mid X)$ and $P(t\mid X)$ capture time-varying vaccine-induced protection against infection and disease, respectively, where $t$ is time since vaccination. Let $\pi(X)$ denote baseline probability of disease given infection in an unvaccinated individual.

**First line: leaky protection against infection**

Vaccination reduces susceptibility to infection by a multiplicative factor

$$S(t\mid X)\in(0,1],$$

so vaccinated infection hazard/risk is

$$\lambda_{V}(t\mid X)=S(t\mid X)\text{ }\lambda_{0}(X).$$

**Second line: leaky protection against progression to disease**

Vaccination reduces disease given infection by a multiplicative factor

$$P(t\mid X)\in(0,1],$$

so

$$\Pr(\text{disease}\mid\text{infection},V,t,X)=P(t\mid X)\text{ }\pi(X).$$

Population-level disease risks

Unvaccinated disease risk

$$R_{SP}^{U}=\mathbb{E}\text{ }\left[ \lambda_{0}(X)\text{ }\pi(X) \right].$$

Vaccinated disease risk (time-varying through $S,P$)

$$R_{SP}^{V}(t)=\mathbb{E}\text{ }\left[ S(t\mid X)\text{ }\lambda_{0}(X)\text{ }P(t\mid X)\text{ }\pi(X) \right].$$

(Expectation is over the population distribution of $X$)

**Vaccine effectiveness estimands**

Effectiveness against infection

$$VE_{S}(t)=1-\frac{\mathbb{E}\text{ }\left[ S(t\mid X)\text{ }\lambda_{0}(X) \right]}{\mathbb{E}\text{ }\left[ \lambda_{0}(X) \right]}.$$

Effectiveness against disease

$$VE_{SP}(t)=1-\frac{\mathbb{E}\text{ }\left[ S(t\mid X)\text{ }\lambda_{0}(X)\text{ }P(t\mid X)\text{ }\pi(X) \right]}{\mathbb{E}\text{ }\left[ \lambda_{0}(X)\text{ }\pi(X) \right]}.$$

**Conditional effectiveness against progression given infection**

The time-varying disease risk among infected:

Vaccinated:

$$\Pr(\text{disease}\mid\text{infection},V,t)=\mathbb{E}\text{ }\left[ P(t\mid X)\text{ }\pi(X)\mid\text{infection},V,t \right].$$

Unvaccinated:

$$\Pr(\text{disease}\mid\text{infection},U)=\mathbb{E}\text{ }\left[ \pi(X)\mid\text{infection},U \right].$$

Then

$$VE_{P}(t)=1-\frac{\mathbb{E}\text{ }\left[ P(t\mid X)\text{ }\pi(X)\mid\text{infection},V,t \right]}{\mathbb{E}\text{ }\left[ \pi(X)\mid\text{infection},U \right]}.$$

$$\mathbb{P}(X\mid\text{infection},V,t)\propto\lambda_{0}(X)\text{ }S(t\mid X),\mathbb{P}(X\mid\text{infection},U)\propto\lambda_{0}(X).$$

So, conditioning on infection reweights the vaccinated infected group by $S(t\mid X)$.

Under this formulation, vaccine effectiveness against infection, VEs(t), reflects the average reduction in infection risk across the population. Vaccine effectiveness against disease, VEsp(t), reflects the average reduction in disease risk and depends jointly on reductions in infection risk and reductions in progression risk. In contrast, vaccine effectiveness against progression given infection, VEp(t), is defined conditionally among infected individuals and therefore averages over a selected, time-varying subset of the vaccinated population. Because individuals with higher residual susceptibility are more likely to experience breakthrough infection, conditioning on infection reweights the vaccinated risk set toward those with weaker protection. When susceptibility to infection is correlated with baseline disease risk or with the durability of progression protection, VEp(t) can vary over time in ways that reflect changing risk-set composition rather than changes in progression biology. By contrast, VEsp(t) and absolute-risk decompositions of disease protection remain stable summaries of vaccine benefit under both leaky and threshold-based protection models.
